# Effectiveness of Ferritin-guided Donation Intervals in Blood Donors: Results of the Stepped-wedge Cluster Randomised FIND’EM Trial

**DOI:** 10.1101/2023.01.24.23284933

**Authors:** Amber Meulenbeld, Steven Ramondt, Maike G. Sweegers, Franke A. Quee, Femmeke J. Prinsze, Emiel O. Hoogendijk, Dorine W. Swinkels, Katja van den Hurk

## Abstract

**Background:** Whole blood donors are at increased risk for iron deficiency and anaemia. The current standard of haemoglobin (Hb) monitoring is insufficient to ensure the maintenance of proper iron reserves and donor health. We determine the effects of ferritin-guided donation intervals for blood donors and the blood supply.

**Methods:** A ferritin-guided donation interval policy was implemented and evaluated through a stepped-wedge cluster-randomised controlled trial in all blood collection centres in the Netherlands. Ferritin was measured in new donors and at every 5th donation. Subsequent donation intervals were extended to six months if ferritin was 15 - 30 ng/mL and to twelve months if ferritin was <15 ng/mL. Primary outcomes are ferritin and Hb levels, iron deficiency, Hb deferral, and donor return. Secondary outcomes are self-reported iron deficiency-related symptoms. Operational statistics assessed the impact on the blood supply.

**Findings:** Among 412,888 whole blood donors, 36,099 were measured. Ferritin-guided intervals increased ferritin levels by 0.24 log10 ng/mL (95% CI 0.22 – 0.27, p <0.001) and Hb by 0.37 g/dL (95% CI 0.31 –0.43, p <0.001). Donors were less likely to be iron deficient (OR 0.15, 95% CI 0.10 – 0.21, p <0.001) or deferred based on Hb (OR 0.23, 95% CI 0.11 – 0.41, p <0.001). However, donors returned less frequently (OR 0.60, 95% CI 0.51 – 0.67, p <0.001), necessitating more invitations and reducing the average donation frequency. No improvement in self-reported symptoms was observed.

**Interpretation:** Ferritin-guided donation intervals significantly improved Hb and ferritin levels, reducing iron deficiency and Hb deferrals. While beneficial for iron and Hb maintenance, efforts are needed to recruit and retain donors and remedy self-reported symptoms.

**Funding and registration:** This project received support from the ’Product and Process Development Cellular Products Grant’ (PPOC18-15) granted to K. van den Hurk by the Research Programming Committee of Sanquin. The trial was registered in the Dutch trial registry (NTR6738) on September 29^th^, 2017 (https://trialsearch.who.int/Trial2.aspx?TrialID=NTR6738).

## Introduction

Annually, more than 100 million whole blood donations are made worldwide to provide life-saving treatment for patients requiring transfusion. Whole blood donors that donate repeatedly are at risk of developing iron deficiency and subsequent iron deficiency anaemia (1). Iron deficiency also has effects beyond that of anaemia since iron is also found in cytochromes and myoglobin. Male donors lose approximately 8% of their total iron stores with each whole blood donation, whereas menstruating women lose on average 81% of their total iron stores (2–4). Iron stores can be monitored through measuring ferritin. Iron deficiency, ferritin levels <15 ng/ml, presents in approximately 15.0% of female and 9.4% of male Dutch repeat donors (5).

There is no consensus on iron management strategies for donor health, but key approaches include extended donation intervals and iron supplementation (6–8). Iron supplementation has been reported to reduce fatigue and benefit exercise performance in iron deficient females (9–11). Concerns about negative perceptions, side effects, and low compliance may favour extended donation intervals over iron supplementation (8,12). In previous studies, extended donation intervals were associated with lower risk of haemoglobin (Hb) deferrals (13,14). The INTERVAL trial showed that longer donation intervals led to significantly higher Hb and ferritin levels (14). Most blood establishments monitor Hb to protect donor health, prevent anaemia and ensure sufficient Hb content in the product, deferring blood donors with Hb levels below 13.5 g/dL (males) or 12.5 g/dL (females) (6–8,15,16). However, iron deficiency can present without anaemia, especially in repeat donors (17). As Hb levels may be within acceptable ranges in the presence of iron deficiency, the current standard is insufficient to maintain proper iron reserves (9). The World Health Organization (WHO) advises additional monitoring of ferritin to detect iron deficiency and monitor donor health.

In 2017, Sanquin Blood Bank in The Netherlands implemented a nationwide policy for iron monitoring of whole blood donors. In addition to Hb monitoring, ferritin is measured at the first visit of new donors and after every fifth whole blood donation of repeat donors (18). If ferritin is between 15 and 30 or below 15 ng/mL, the subsequent donation interval is extended to six or twelve months, respectively (18). A stepped-wedge approach was used to evaluate the effects of this ferritin-guided donation interval policy for whole blood donors on (1) Hb and ferritin levels, (2) iron deficiency, (3) Hb deferral, (4) donor return rates, (5) self-reported iron deficiency-related symptoms, and (6) the blood supply.

## Methods

### Design

Ferritin measurement IN Donors—Effectiveness of iron Monitoring (FIND’EM) is a stepped-wedge cluster randomised trial, described in detail by Sweegers et al. (18). We conducted cross-sectional ferritin measurements for all donors visiting a blood collection centre during pre-selected weeks before, during, and after implementing a new policy. We measured ferritin in stored plasma samples from donors who consented to use their leftover materials for medical research. Additionally, optional questionnaires were offered to donors. The trial was powered on 4ng/mL change in the ferritin level outcome over six months, assuming a SD of 54 ng/mL, ICC of 0.10 and 150 visits per donation centre (18). The policy and implementation strategy were approved by Sanquin’s Medical and Ethical Advisory Board. The trial was approved by Sanquin’s Board of Directors.

### Randomization

The 138 fixed and mobile blood collection centres in the Netherlands are divided over 29 staffing clusters that we randomly assigned to four randomization groups. The stepped-wedge design involved random and sequential rollout of ferritin-guided donation intervals by randomization group from November 2017 to November 2019. The allocation was not concealed and the timing of the rollout was revealed before the study. Donors were notified of the new policy through mail upon implementation at their regular donation centre.

### Procedures

As part of routine procedures, donors undergo a health screening to assess their eligibility to donate by measuring Hb levels and blood pressure and assessing recent travel, sexual and medical history prior to every donation. In the Netherlands, donors also make a screening visit prior to their first donation. In accordance with European legislation, Hb cut-offs for donor eligibility are >13.5 g/dL for males and >12.5 g/dL for females. Donors with low Hb (capillary measurement prior to donation with HemoCue 201, Angelholm, Sweden) are deferred for 3 months. If eligible, males can donate every 56 days up to five times a year, females every 122 days up to three times a year, independent of menopausal status.

The intervention entails extended donation intervals based on ferritin measurements. The cut-offs for extended intervals were based on definitions for iron deficiency (<15 ng/mL, 12 months deferral) and reduced iron levels (≤30 ng/mL, 6 months deferral) by Alvarez-Ossorio et al. (8). The six month interval is based on the results from Schotten et al., suggesting intervals of at least 180 days are required to fully replete iron stores after donation (7). A twelve-month interval is indicated for iron deficient donors to replete to higher levels than pre-donation. In whole blood donors, ferritin is measured at the pre-donation screening visit before the first donation and of each fifth whole blood donation. If ferritin levels are >30 ng/mL, donors are eligible to donate at regular donation intervals. If ferritin levels are ≤30 ng/mL but ≥15 ng/mL, repeat donors are deferred for six months; new donors are allowed to donate, but ferritin is measured again at this first donation, after which they follow the repeat donor policy (18). If ferritin levels are <15 ng/mL, new and repeat donors are deferred from donating for twelve months. Ferritin levels are measured again in the donation after this extended interval. Donor physicians may additionally advise donors to visit their general practitioner and/or donate at a lower frequency (18).

### Outcome assessment

Before the trial, we established four measurement weeks: 1) one week before implementation (baseline), 2) midway through implementation, 3) one week after full implementation, and 4) one year after full implementation of ferritin-guided donation intervals. Primary outcomes included Hb and ferritin levels, Hb deferral, iron deficiency, and donor return rates, while secondary outcomes included iron deficiency-related symptoms and the impact on the blood supply. Pseudonymized data on age, sex, Hb levels, and deferrals were extracted from the blood bank database (eProgesa software application; MAK-SYSTEM, Paris, France) for donors who visited a blood collection centre during a measurement week and consented to their leftover materials being used for research. Donor return was defined as attempting a donation within six months after becoming eligible to donate again, accounting for minimum donation intervals and possible deferrals. We measured ferritin levels on the Architect i2000sr (Abbott Diagnostics) in plasma samples collected at donation in K3EDTA tubes (VACUETTE®, Greiner Bio-One, Kremsmünster, Austria) and stored at -30⁰C for three to 24 months (19). The ferritin assay used is traceable to the first WHO Human Liver Ferritin International Standard (80/602). Between-day precision (CV%) of the ferritin assay is 4.1% (95% CI 3.8-4.3%) and constant over a concentration range of 10-500 ng/mL.

New and repeat donors in fixed blood collection centres during measurement week 2 and 3 that provided informed consent were asked to complete questionnaires. These questionnaires assess physical and mental wellbeing (SF-36), fatigue (Checklist Individual Strength), donation-related symptoms, restless legs syndrome (RLS) and pica, cognitive functioning (Cognitive Failure Questionnaire), and warm glow.(18,20–23) Questionnaires were completed using Qualtrics XM® software (Provo, UT, USA) during or shortly after the donation (18).

We aimed to assess the impact of the new policy on the blood supply by comparing operational data from the blood bank before, during and after the study period. We gathered data on whole blood donations, donors and donations invitations from the blood bank database (eProgesa software application; MAK-SYSTEM, Paris, France). Available donors were defined as not being deferred for medical (iron-related or other), travel or lifestyle factors; new donors as showing up for the pre-donation screening; and lapsed donors as deregistered or unresponsive to 5 subsequent invitations. Demands for each year were based on hospital requests and converted to blood units.

### Data analyses

Descriptive statistics are provided for the donors measured during measurement week 1. Using donors measured during all four weeks, we conducted complete case analysis and used ordinary least squares regression in case of continuous outcomes and logistic regression for binary outcomes. After diagnostic tests of linearity and errors assumptions, a log base 10 transformation was applied to improve linearity of ferritin. We conducted robust regression to analyse cognitive functioning, physical health, mental health and fatigue, as the normality assumption for logistic regression was violated. Measurements after implementation of the new policy were grouped in periods of 6 months and compared to the control condition. The control condition was defined as no implementation or implementation less than 6 months ago. Analyses were adjusted for age, weight and height, and stratified by sex and menopausal status. Donor return analyses were adjusted for return being during a COVID-19 lockdown. Null hypotheses are that there are no associations between implementation of the new policy and the outcomes. A two-sided p-value of <0.05 is considered statistically significant and a Bonferroni correction is applied. Statistical analyses were conducted using the R language and environment for statistical computing (4.1.1) and the lme4 (1.1.33), car (3.1.2), lmtest (0.9.40), sandwich (3.0.2), olsrr (0.5.3), and robustbase (0.95.1) packages. HTML files of the scripts and results produced in Quarto are available through https://github.com/Sanquin/FINDEM.

### Deviations from protocol

As the number of donors with low ferritin in the first implementation group was higher than anticipated, further implementation was executed in smaller steps to mitigate the impact on the blood supply (18). A fourth measurement week was added, a year after the third measurement week, between November 23^rd^ and 27^th^ 2020). This fourth week enables evaluation of long-term results. We furthermore deviated from the protocol by using ordinary least squares (OLS) regression instead of linear mixed models. In the linear mixed models for analysis of the primary outcomes with random effects for the donation centre clusters, the ICC ranged from 0.00 to 0.05; R^2^ did not improve with random effects; and residual variance exceeded the variance between clusters. This indicates that the random effects of the donation centre clusters were of minimal importance and justifies the use of OLS regression. We conducted sensitivity analyses for donor return, stratifying by ferritin level, reason for deferral, and donation centre type. Lastly, we added a sixth element (impact on the blood supply) to the research question.

## Results

### Donor characteristics

412,888 whole blood donors made 1,634,700 donations during the study period. We performed measurements in samples of 37,621 donations by 36,099 donors (Figure 1). Overall, across the randomization groups, baseline donor characteristics were well balanced and comparable with the general donor population (Table 1, Table S1). Characteristics of the 7,573 participants in the questionnaire are comparable to the baseline participants (Table S2).

**Figure 1.**
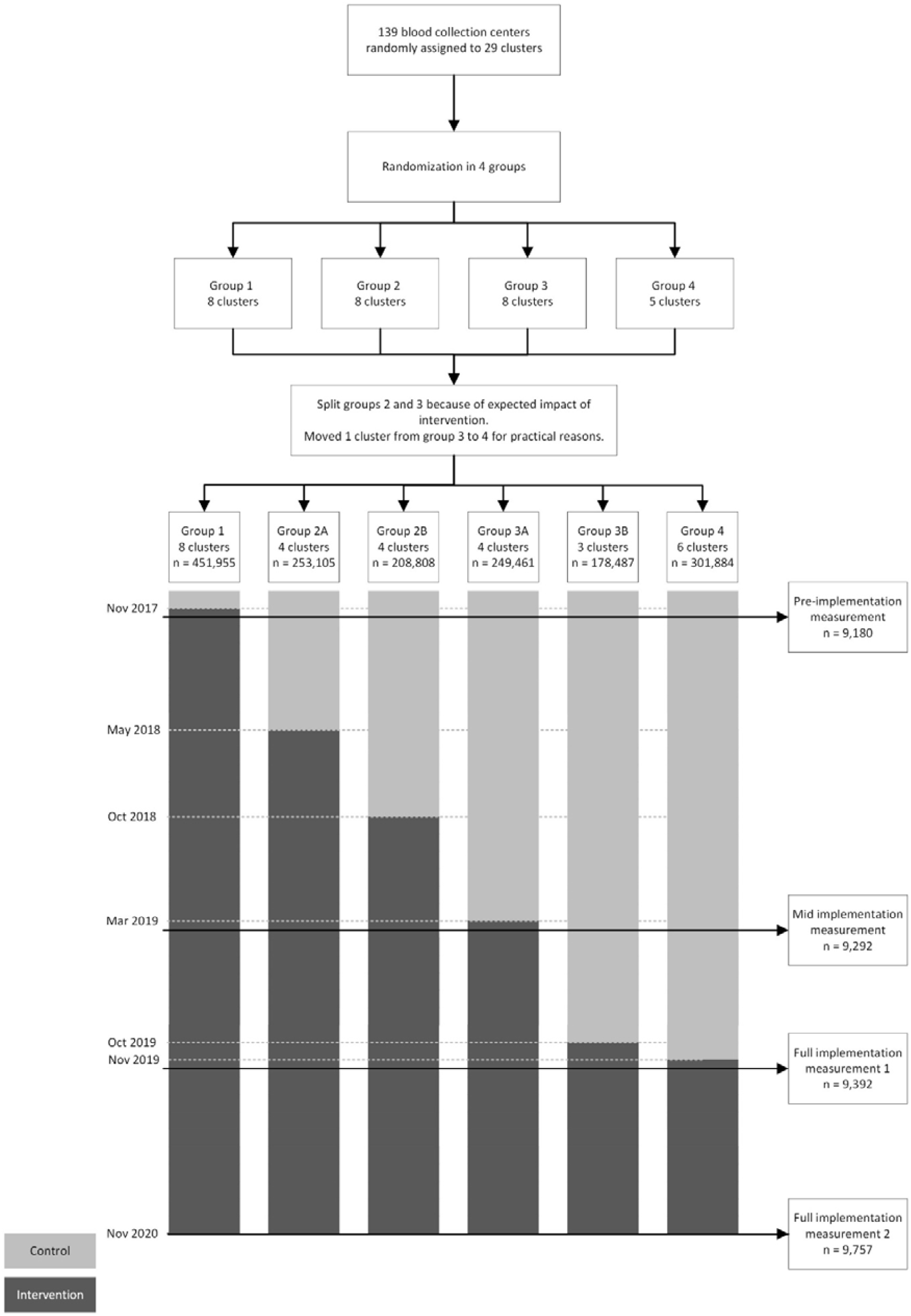
Flow chart of stepped-wedge cluster randomized trial and intervention roll out by randomization group

**Table 1.** Donor characteristics in measurement week 1, where group 1 had just implemented the new policy and groups 2, 3, and 4 were considered controls.

|  | Group 1<sup>1</sup> | Group 2 | Group 3 | Group 4 | Overall |
| --- | --- | --- | --- | --- | --- |
| Pre-menopausal, female donors | N = 688 | N = 590 | N = 634 | N = 465 | N = 2,377 |
| New donors, N (%) | 167 (24.3) | 109 (18.5) | 131 (20.7) | 86 (18.5) | 493 (20.7) |
| Age (years), median [IQR] | 25.0 [21.0, 33.0] | 26.0 [21.0-34.0] | 26.0 [22.0-33.8] | 25.0 [21.0-31.0] | 25.0 [21.0-33.0] |
| Height (cm), mean (SD) | 171 (6.4)* | 171 (6.9) | 171 (6.3)* | 171 (6.4) | 171 (6.5)* |
| Weight (kg), mean (SD) | 71.0 (12.2)* | 71.0 (12.4) | 71.4 (11.9) | 70.8 (12.1) | 71.1 (12.2)* |
| Hemoglobin, median [IQR], g/dL | 13.4 [12.7-12.4] | 13.4 [12.9-14.0] | 13.5 [12.9-14.0] | 13.4 [12.9-14.0] | 13.4 [12.9-14.0] |
| Low hemoglobin deferral (<12.5 g/dL), N (%) | 64 (9.3) | 34 (5.8) | 46 (7.3) | 43 (9.2) | 187 (7.9) |
| Ferritin, median [IQR], ng/mL | 26.2 [13.3-46.2]** | 24.4 [12.1-46.7]** | 24.7 [12.7-45.5]** | 23.7 [10.3-40.3]** | 25.1 [12.3-45.4]** |
| Iron deficiency (ferritin < 15 ng/mL), N (%) | 161 (23.4)** | 159 (26.9)** | 173 (27.3)** | 130 (28.0)** | 623 (26.2)** |
| Total number of whole blood donations, median [IQR] | 2.0 [1.0-7.0] | 3.0 [1.0-9.0] | 4.0 [1.0-10.0] | 3.0 [1.0-9.0] | 3.0 [1.0-9.0] |
| Number of whole blood donations in the past 2 years, median [IQR] | 2.0 [1.0-4.0] | 2.0 [1.0-4.0] | 2.0 [1.0-4.0] | 3.0 [1.0-4.0] | 2.0 [1.0-4.0] |
| Donor return within 6 months of first allowed date, N (%) | 478 (66.5) | 383 (64.9) | 436 (68.8) | 323 (69.5) | 1,620 (68.2) |
| Post-menopausal, female donors | N = 388 | N = 379 | N = 387 | N = 308 | N = 1,462 |
| New donors, N (%) | 11 (2.8) | 7 (1.8) | 13 (3.4) | 2 (0.6) | 33 (2.3) |
| Age (years), median [IQR] | 55.0 [49.0-60.0] | 55.0 [50.0-61.0] | 56.0 [49.0-60.0] | 55.0 [49.8-62.0] | 55.0 [50.0-61.0] |
| Height (cm), mean (SD) | 170 (5.9) | 169 (6.1) | 170 (6.2) | 170 (5.9) | 170 (6.0) |
| Weight (kg), mean (SD) | 74.5 (11.3) | 73.4 (12.3) | 73.9 (12.4) | 73.3 (11.6) | 73.8 (11.9) |
| Hemoglobin, median [IQR], g/dL | 13.9 [13.1-14.5] | 13.5 [13.1-14.5] | 13.7 [13.1-14.3] | 13.5 [13.1-14.3] | 13.7 [13.1-14.3] |
| Low hemoglobin deferral (<12.5 g/dL), N (%) | 19 (4.9) | 19 (5.0) | 21 (5.4) | 11 (3.6) | 70 (4.8) |
| Ferritin, median [IQR], ng/mL | 27.4 [13.5-44.0]*** | 28.3 [14.0-45.8]** | 25.7 [13.2-45.0]** | 26.7 [15.0-43.6]** | 27.4 [13.7-45.0]** |
| Iron deficiency (ferritin < 15 ng/mL), N (%) | 85 (21.9)*** | 87 (23.0)** | 95 (24.5)** | 64 (20.8)** | 331 (22.6)** |
| Total number of whole blood donations, median [IQR] | 29.0 [11.0-50.0] | 26.0 [12.0-42.0] | 29.0 [12.0-46.0] | 29.0 [12.0-47.0] | 28.0 [12.0-46.0] |
| Number of whole blood donations in the past 2 years, median [IQR] | 5.0 [3.0-5.0] | 4.0 [3.0-5.0] | 5.0 [3.0-6.0] | 5.0 [3.0-5.0] | 5.0 [3.0-6.0] |
| Donor return within 6 months of first allowed date, N (%) | 297 (76.5) | 292 (77.0) | 310 (80.1) | 237 (76.9) | 1,136 (77.7) |
| Male donors | N = 1075 | N = 1061 | N = 1007 | N = 870 | N = 4,013 |
| New donors, N (%) | 58 (5.4) | 63 (5.9) | 46 (4.6) | 47 (5.4) | 214 (5.3) |
| Age (years), median [IQR] | 48.0 [32.0-58.5] | 51.0 [35.0-60.0] | 49.0 [32.5-58.0] | 49.0 [31.0-59.0] | 49.0 [32.0-59.0] |
| Height (cm), mean (SD) | 184 (7.1) | 183 (7.2) | 183 (6.9) | 183 (7.5) | 183 (7.2) |
| Weight (kg), mean (SD) | 86.5 (12.1) | 85.8 (12.2) | 85.4 (12.0) | 85.4 (11.9)* | 85.8 (12.1) |
| Hemoglobin, median [IQR], g/dL | 14.8 [14.2-15.6] | 14.8 [14.2-15.6] | 14.8 [14.2-15.6] | 14.8 [14.2-15.5] | 14.8 [14.2-15.6] |
| Low hemoglobin deferral (<13.5 g/dL), N (%) | 42 (3.9) | 27 (2.5) | 34 (3.4) | 27 (3.1) | 130 (3.2) |
| Ferritin, median [IQR], ng/mL | 25.3 [12.3-54.9]** | 25.9 [13.2-55.3]** | 24.8 [12.4-50.0]** | 24.5 [11.6-54.8]** | 25.2 [12.4-53.2]** |
| Iron deficiency (ferritin < 15 ng/mL), N (%) | 280 (26.0)** | 273 (25.7)** | 272 (27.0)** | 243 (27.9)** | 1,068 (26.6)** |
| Total number of whole blood donations, median [IQR] | 30.0 [7.0-68.0] | 39.0 [8.0-71.0] | 32.0 [8.0-64.0] | 31.5 [7.0-70.0] | 32.0 [7.0-68.0] |
| Number of whole blood donations in the past 2 years, median [IQR] | 6.0 [4.0-9.0] | 6.0 [4.0-9.0] | 6.0 [4.0-8.0] | 6.0 [4.0-8.0] | 6.0 [4.0-8.0] |
| Donor return within 6 months of first allowed date, N (%) | 924 (86.0) | 905 (85.3) | 870 (86.4) | 738 (84.8) | 3,437 (85.6) |

<sup>1</sup>Randomly assigned clusters of donation centres (Group 1-3: 8 clusters, group 4: 5 clusters)
Missing: \* < 10%, \*\* 10-20%, \*\*\* > 20%

### Ferritin, iron deficiency, Hb, and Hb deferral

The prevalence of iron deficiency and Hb deferral decreased after implementation of the policy (Figure 2). Ferritin-guided donation intervals are associated with significantly higher ferritin and Hb levels and less iron deficiency and Hb deferrals (Figure 3a). Compared to pre-implementation, the intervention was associated with up to 0.24 ng/mL higher log10 ferritin and 0.37 g/dL higher Hb at 36 months after implementation in males (Figure 3a). In premenopausal and postmenopausal females, we also observe positive, but smaller, associations with an increasing trend over time. Moreover, the odds ratio of 0.19 indicates that males are 81% less likely to be iron deficient at 36 months after implementation than before the intervention (Figure 3a). In postmenopausal females, odds ratios also decrease over the course of the trial, with iron deficiency being 87% less likely at 36 months after implementation compared to pre-implementation (Figure 3a). Hb deferral in males was up to 77% less likely as time since implementation increased, compared to pre-implementation. Similarly, in premenopausal females, Hb deferral is up to 44% less likely at 30-35 months after implementation than at pre-implementation (Figure 3a). In postmenopausal females, Hb deferral is up to 51% less likely at 36 months after implementation than at pre-implementation.

**Figure 2.**
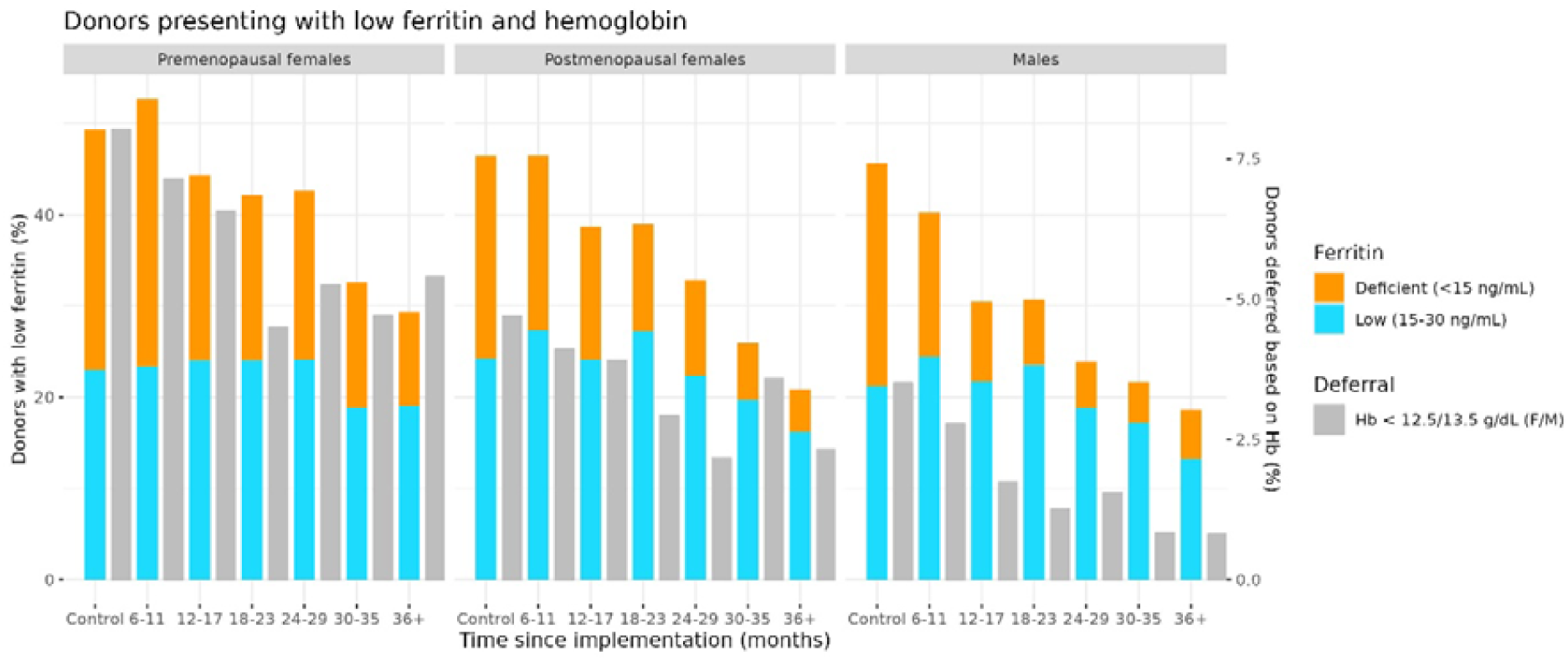
Percentage of low Hb and ferritin deferral per sex in controls and at six timepoints post-intervention.

**Figure 3.**
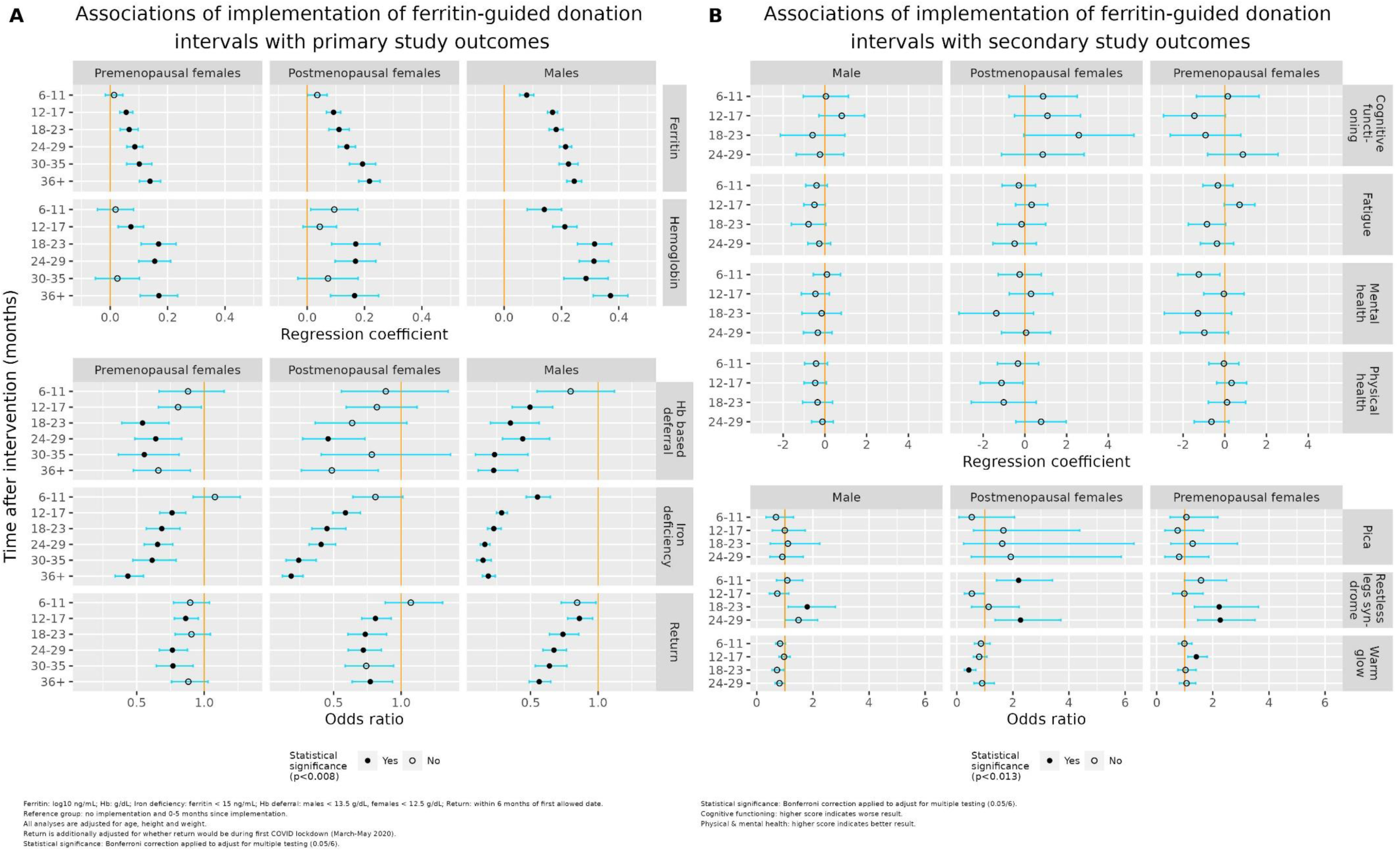
Results of regression analyses for iron-related outcomes, donor return and iron-deficiency related symptoms. Corresponding coefficients, confidence intervals and p-values are displayed in Table S3.

### Donor return

After adjustment for return being during a COVID-19 lockdown, odds for donor return within six months are lower at each time point after implementation compared to pre-implementation (Figure 3a). Generally, the negative association with donor return was stronger in males. At 36 months after implementation, males were 42% less likely to return for donation (Figure 3a). For premenopausal and postmenopausal females, we do not observe a consistent decrease in odds over time (Figure 3a). The differences in odds between groups of ferritin levels, reasons for deferral and donation centre types are mostly not statistically significant (Table S4-6).

### Iron deficiency-related symptoms

There were no consistent significant patterns for pica, fatigue, cognitive functioning, the SF-36, and warm glow after implementation of the new policy (Figure 3b). However, males and premenopausal and postmenopausal females were 49%, 127%, and 127% more likely to report RLS at 24 months after implementation than before implementation.

### Donor deferral, donation frequency and blood supply

Over time after implementation of the policy, the number of donations decreased in line with decreasing demand, but the number of invitations needed to obtain one donation increased (Table 2). The average donation frequency of donors decreased over the years and subsequently the number of donors that donated increased. The number of available donors increased and the proportion that donated fluctuated between 80 and 90% over the years. The number of simultaneously unavailable donors stabilized at around 40,000 donors per year. While the percentage of low ferritin deferrals increased as a result of the implemented policy, the percentage of low-Hb deferrals decreased. The number of new donors at the blood bank was higher in 2019 and 2020 than in other years, while the number of lapsed donors in 2019 was exceptionally low.

**Table 2.** Blood bank statistics per calendar year to describe impact of new policy on blood supply in the Netherlands. Ferritin-guided donation intervals were implemented, in a step-wise manner, in November 2017. Hence, there is no data on extended intervals due to low ferritin in 2016. In November 2019 implementation was completed and in November 2020 the study was completed. All statistics are presented for whole blood donations only.

|  | 2016 | 2017 | 2018 | 2019 | 2020 | 2021 | 2022 |
| --- | --- | --- | --- | --- | --- | --- | --- |
| Hospital demand (in donation units) | 418,491 | 407,215 | 406,199 | 407,460 | 402,578 | 400,757 | 392,805 |
| Donations | 421,019 | 410,956 | 412,266 | 412,479 | 411,021 | 406,845 | 393,633 |
| Donors | 234,821 | 234,224 | 237,985 | 248,911 | 278,863 | 262,956 | 260,033 |
| Average donation frequency per donor | 1.79 | 1.75 | 1.73 | 1.66 | 1.47 | 1.55 | 1.51 |
| Average invitations per donation | 2.83 | 2.95 | 3.03 | 3.12 | 3.19 | 3.41 | 3.59 |
| Extended donation intervals<sup>1</sup> |  |  |  |  |  |  |  |
| Number of donations | - | 1,979 | 19,044 | 45,829 | 60,938 | 63,695 | 66,635 |
| Percentage of donations | - | 0.48 | 4.62 | 11.11 | 14.83 | 15.66 | 16.93 |
| Unique donors | - | 1,973 | 17,441 | 38,961 | 53,841 | 51,140 | 45,696 |
| Hb deferral |  |  |  |  |  |  |  |
| Number of donations | 38,143 | 39,083 | 37,733 | 30,411 | 20,479 | 21,589 | 21,494 |
| Percentage of donations | 9.06 | 9.51 | 9.15 | 7.37 | 4.98 | 5.31 | 5.46 |
| Unique donors | 32,008 | 32,844 | 31,539 | 26,222 | 20,599 | 16,813 | 17,820 |
| Available donors<sup>2</sup> | 286,597 | 273,379 | 283,550 | 305,225 | 312,930 | 308,751 | 322,637 |
| Unavailable donors due to low ferritin | - | 1,973 | 14,435 | 32,585 | 42,709 | 39,570 | 38,901 |
| Available donors invited to donate (%) | 81.93 | 85.68 | 83.93 | 81.55 | 80.11 | 85.17 | 80.60 |
| New donors | 50,107 | 44,362 | 58,419 | 60,121 | 71,983 | 54,194 | 53,194 |
| Lapsed donors | 49,066 | 51,579 | 42,782 | 26,571 | 47,918 | 55,341 | 29,982 |

<sup>1</sup> Due to low ferritin (including both <15 ng/mL and 15-30 ng/mL)
<sup>2</sup> On December 31

## Discussion

This stepped-wedge cluster-randomised trial in whole blood donors shows that ferritin and Hb levels are significantly higher and iron deficiency and Hb deferral significantly lower after the implementation of ferritin-guided donation intervals compared to the prior standard of Hb monitoring only. Conversely, we found that the odds of donor return within six months decreased substantially with this policy. Most iron deficiency-related symptoms remained unaffected, except for an increase in reports of RLS. The continuity of the blood supply was maintained, but additional efforts were required to account for the decreased donor availability.

Ferritin-guided donation intervals were associated with significantly less iron deficiency and higher ferritin and Hb levels compared to pre-implementation, consistent with findings from simulations assessing the impact of ferritin testing (24). The weaker effects observed in premenopausal females could be due to their already less frequent donations before the policy, driven by longer donation intervals and more frequent low-Hb deferrals, providing more recovery time (25,26). Additionally, because of more stable iron stores due to the healthy donor effect, combined with lower donation frequency and ferritin levels in this group, may result in less pronounced improvements compared to other groups (19,27). Our finding that the new policy is associated with significantly lower odds of donor return supports previous findings on decreased return rates after deferral (26,28). The weaker negative association with donor return in premenopausal females compared to other groups may again be explained by their exposure to more frequent deferral through the old Hb monitoring policy.

The increased reporting of RLS seems surprising but may simply be a result of increased awareness through the letter sent to donors upon implementation in their donation centre, in which restless legs syndrome was specifically mentioned as a symptom of iron deficiency. Further investigation shows no statistically significant correlation between RLS and ferritin levels. The lack of improvement we found in self-reported iron deficiency-related symptoms may partially be due to the small number of questionnaire participants before implementation. However, our findings align with previous randomised placebo-controlled trials investigating iron repletion and self-reported donor health (29–31). Moreover, the benevolence and warm glow that drive donors may overshadow negative health consequences which stresses the need for more functional and objective measures of iron deficiency-related symptoms like fatigue (23). Positive effects of the steady reduction in iron deficiency and Hb deferral on self-reported iron deficiency-related symptoms may also be delayed and require longer follow-up in future studies.

Given the results of this trial regarding donor health and the continued fulfilment of the hospital demand during and after the implementation of ferritin-guided donation intervals, Sanquin Blood Bank decided to keep the policy in place. Our findings do, however, raise important policy and practical considerations. First, it provides policymakers with a monitoring option that requires no effort from blood donors, may be more effective in protecting iron status than standard Hb monitoring practices, and does not impose a risk of side effects compared to iron supplementation (12). However, the decreasing donor availability due to decreasing odds of donor return and increased numbers of deferred donors stress the need for intensified donor recruitment and retention efforts (25). This is reflected in the blood bank practice, as more invitations are needed to obtain one donation. Moreover, as donation frequencies decreased after implementation of the policy, more donors were needed to reach donation demands. To increase donor availability, iron supplementation can be considered as an alternative or additional strategy (10,24).

To our knowledge, this is the first randomised study that goes beyond the limitations of Hb monitoring by evaluating the effectiveness of ferritin-guided donation intervals in improving Hb and ferritin levels, donor return rates and decreasing iron deficiency, Hb deferrals, iron deficiency-related symptoms. One of the major strengths is the stepped-wedge cluster randomised approach as it allows simultaneous implementation and evaluation of a new policy. However, there are also limitations to this study. First, the nature of the intervention poses a risk of overestimating the positive effect of ferritin-guided donation intervals. Donors whose donation intervals are extended, may return less frequently, potentially resulting in selection bias. The risk of overestimation is to some extent minimized as the duration of the trial exceeds the length of extended donation intervals. There may also be undetected delayed effects in this trial, because ferritin levels are measured at every fifth donation, and thus individual donors may not be confronted with the policy immediately upon implementation in their donation centre.

The FIND’EM study provides substantial evidence that ferritin-guided donation intervals increase Hb and ferritin levels and thereby effectively prevent iron deficiency and Hb deferral, while efforts are needed to mitigate effects on the blood supply. Future studies may investigate long-term effects on donor health outcomes. Our results may help blood services worldwide to improve their donor iron management policies in order to protect donors from developing iron deficiency.

## Declarations

### Ethics approval and consent to participate

Sanquin’s Ethical Advisory Board has reviewed and approved the FIND’EM protocol. Only data from donors who gave consent for their information and left-over material to be used for research purposes at the health check were included in the study. Donors who completed questionnaires provided informed consent for their questionnaire data to be matched with data from the donor database.

## Declaration of interests

The authors declare that they have no competing interests.

## Funding

The current project is supported by the ‘Product and Process Development Cellular Products Grant’ (PPOC18-15) granted to K. van den Hurk by the Research Programming Committee, Sanquin, Amsterdam, The Netherlands.

## Authors’ contributions

KvdH designed the study. KvdH, MS, FP and FQ set up and coordinated data collection. AM, and SR wrote the analysis plan and drafted the manuscript. AM, SR and FP conducted the analyses with support from EH. The manuscript was critically reviewed and approved by all co-authors.

## Data sharing

The study protocol of the FIND’EM trial including a statistical analysis plan was previously published. Analysis code used to produce the results reported in this article are available online. Individual participant data, after de-identification, that underlie the results reported in this article (text, tables, figures, and appendices) are available to researchers from GDPR compliant institutions, after provision of a methodologically sound and feasible proposal. Requests can be submitted to the authors immediately following publication. A material transfer or research collaboration agreement has to be agreed and signed with the researcher.

## Acknowledgements

We would like to acknowledge all blood bank staff for their commitment to the implementation of the policy and their efforts in collecting data for our study. We are thankful to all donors for their contribution to the study and the blood supply.

## Notes

### Competing Interest Statement

The authors have declared no competing interest.

### Clinical Trial

NTR6738

### Clinical Protocols

https://pubmed.ncbi.nlm.nih.gov/32998766/

### Author Declarations

The Board of Directors, the Medical Advisory Council and Ethics Advisory Council of Sanquin Blood Supply Foundation gave ethical approval for this work.

### Summary of Updates

This manuscript has been revised to include additional analyses on the impact of the implemented policy on the national blood supply (presented in Table 2). The abstract has been updated to also include these analyses.

