## Supplemental table 1 for "Effectiveness of Ferritin-guided Donation Intervals in Blood Donors: Results of the Stepped-wedge Cluster Randomised FIND’EM Trial"

| **Table S1.** Donor characteristics of the donors measured in measurement week 1 (before implementation) and measurement week 4 (after full implementation). | |  |  |
| --- | --- | --- | --- |
|  | Week 1^1^ | | Week 4 |
| Pre-menopausal, female donors | N = 2,695 | | N = 3,213 |
| Age (years), median [IQR] | 26.0 [21.0, 34.0] | | 27.0 [22.0, 34.0] |
| Height (cm), mean (SD) | 171 (6.75)* | | 171 (6.49)* |
| Weight (kg), mean (SD) | 71.3 (12.3)* | | 70.8 (16.8)* |
| Hemoglobin, median [IQR], g/dL | 13.4 [12.9, 14.0] | | 13.5 [12.9, 14.2] |
| Low hemoglobin deferral (<12.5 g/dL), N (%) | 64 (9.3) | | 43 (9.2) |
| Ferritin, median [IQR], ng/mL | 25.4 [12.3, 46.1]** | | 29.8 [17.2, 48.7]*** |
| Iron deficiency (ferritin < 15 ng/mL), N (%) | 697 (25.9)** | | 419 (13.0)*** |
| Donor return within 6 months of first allowed date, N (%) | 1769 (65.6) | | 1915 (59.6) |
| Post-menopausal, female donors | N = 1,825 | | N = 2,068 |
| Age (years), median [IQR] | 55.0 [50.0, 61.0] | | 55.0 [50.0, 62.0] |
| Height (cm), mean (SD) | 169 (6.01)* | | 170 (6.48)* |
| Weight (kg), mean (SD) | 73.9 (11.9) | | 74.2 (17.5)* |
| Hemoglobin, median [IQR], g/dL | 13.7 [13.1, 14.5] | | 13.9 [13.2, 14.5] |
| Low hemoglobin deferral (<12.5 g/dL), N (%) | 82 (4.5) | | 61 (2.9) |
| Ferritin, median [IQR], ng/mL | 28.4 [14.3, 46.6]** | | 37.2 [24.8, 58.5]*** |
| Iron deficiency (ferritin < 15 ng/mL), N (%) | 395 (21.6)** | | 157 (7.6)*** |
| Donor return within 6 months of first allowed date, N (%) | 1320 (72.3) | | 1399 (67.7) |
| Male donors | N = 4,660 | | N = 4,476 |
| Age (years), median [IQR] | 50.0 [34.0, 59.0] | | 47.0 [30.0, 59.0] |
| Height (cm), mean (SD) | 183 (7.23) | | 183 (7.11) |
| Weight (kg), mean (SD) | 85.9 (12.1)* | | 85.6 (12.5) |
| Hemoglobin, median [IQR], g/dL | 14.8 [14.2, 15.6] | | 15.3 [14.5, 16.0] |
| Low hemoglobin deferral (<13.5 g/dL), N (%) | 146 (3.1) | | 50 (1.1) |
| Ferritin, median [IQR], ng/mL | 26.7 [13.2, 55.7]** | | 44.2 [27.9, 75.1]** |
| Iron deficiency (ferritin < 15 ng/mL), N (%) | 1167 (25.0)** | | 264 (5.9)** |
| Donor return within 6 months of first allowed date, N (%) | 3875 (83.2) | | 3481 (77.8) |
| ^1^Randomly assigned clusters of donation centres (Group 1-3: 8 clusters, group 4: 5 clusters)  Missing: * < 10%, ** 10-20%, *** > 20% | | | |
