## Supplemental table 2 for "Effectiveness of Ferritin-guided Donation Intervals in Blood Donors: Results of the Stepped-wedge Cluster Randomised FIND’EM Trial"

| **Table S2.** Donor characteristics questionnaire at measurement week 2 | | | | | |
| --- | --- | --- | --- | --- | --- |
|  | Step 1 | Step 2 | Step 3 | Step 4 | Overall |
| Premenopausal, female donors | N = 656 | N = 675 | N = 657 | N = 460 | N = 2,448 |
| Age, median [IQR] | 27.0 [23.0 - 34.0] | 26.0 [22.0 - 34.0] | 26.0 [22.0 - 34.0] | 26.0 [22.0 - 33.0] | 26.0 [22.0 - 34.0] |
| Ferritin, median [IQR] | 24.2 [13.5 - 43.5]* | 22.6 [12.0 - 38.5]* | 20.6 [11.7 - 38.8]* | 22.0 [ 11.1 - 38.8]* | 22.7 [12.1 - 39.9]* |
| Restless legs syndrome, N (%) | 41 (6.3)** | 50 (7.4)* | 51 (7.8)** | 29 (6.3)* | 171 (7.0)** |
| Pica, N (%) | 11 (1.8)* | 13 (2.0)* | 12 (1.9)* | 18 (4.0)* | 54 (2.3)* |
| Fatigue^1^, mean (SD) | 82.3 (6.5)* | 82.5 (6.5)* | 82.4 (6.2)* | 81.8 (6.2)* | 82.3 (6.4)* |
| Cognitive functioning^1^, mean (SD) | 31.7 (14.0)* | 29.4 (12.6)* | 31.3 (12.8)* | 32.1 (13.5)* | 31.0 (13.3)* |
| Physical health (SF-36)^2^, mean (SD) | 89.5 (9.6)* | 90.3 (9.9)* | 89.4 (10.6)* | 89.7 (9.3)* | 89.7 (9.9)* |
| Mental health (SF-36)^2^, mean (SD) | 76.6 (14.1)* | 76.6 (15.1)* | 76.4 (14.9)* | 74.4 (16.5)* | 76.1 (15.1)* |
| Warm glow, N (%) | 312 (55.4)** | 341 (56.6)** | 304 (51.9)** | 193 (46.4)* | 1150 (53.0)** |
| Postmenopausal, female donors | N = 352 | N = 379 | N = 456 | N = 304 | N = 1,491 |
| Age, median [IQR] | 55.0 [50.0 - 61.0] | 55.0 [50.0 - 60.5] | 55.0 - [61.0] | 55.0 [50.0 - 61.2] | 55.0 [ 50.0 - 61.0] |
| Ferritin, median [IQR] | 29.5 [17.7 - 43.1]* | 27.1 [16.5 - 44.3]* | 25.4 [13.4 - 43.7]* | 26.1 [14.9 - 44.0]* | 27.0 [15.3 - 44.0]* |
| Restless legs syndrome, N (%) | 28 (8.0)** | 27 (7.1)** | 60 (13.2)** | 35 (11.5)** | 150 (10.1)** |
| Pica, N (%) | 8 (2.3)* | 6 (1.6)* | 6 (1.3)* | 5 (1.7)* | 25 (1.7)* |
| Fatigue^1^, mean (SD) | 85.9 (7.0)* | 86.0 (6.6)* | 86.4 (6.5)* | 85.4 (5.9)* | 86.0 (6.5)* |
| Cognitive functioning^1^, mean (SD) | 26.5 (11.8)* | 26.4 (11.1.)* | 25.6 (10.3)* | 26.4 (12.1)* | 26.2 (11.2)* |
| Physical health (SF-36)^2^, mean (SD) | 88.4 (11.7)* | 87.6 (12.2)* | 88.9 (10.5)* | 88.9 (10.0)* | 88.5 (11.2)* |
| Mental health (SF-36)^2^, mean (SD) | 81.4 (12.6)* | 81.1 (11.4)* | 81.3 (11.7)* | 80.5 (13.0)* | 81.1 (12.1)* |
| Warm glow, N (%) | 209 (62.8)* | 223 (63,7)* | 290 (67.4)* | 188 (64.6)* | 910 (64.8)* |
| Male donors | N = 848 | N = 925 | N = 1073 | N = 788 | N = 3634 |
| Age, median [IQR] | 47.0 [32.0 - 58.0] | 47.0 [31.0 - 59.0] | 47.0 [32.0 - 58.0] | 49.0 [32.0 - 60.0] | 48.0 [32.0 - 59.0] |
| Ferritin, median [IQR] | 39.3 [24.4 - 68.7]* | 34.0 [18.9 - 59.6]* | 27.8 [13.8 - 51.9]* | 25.8 [12.6 - 52.1]* | 32.0 [16.9 - 57.7]* |
| Restless legs syndrome, N (%) | 44 (5.2)* | 48 (5.2)* | 61 (5.7)** | 61 (7.7)* | 214 (5.9)* |
| Pica, N (%) | 22 (2.6)* | 24 (2.7)* | 27 (2.6)* | 22 (2.9)* | 95 (2.7)* |
| Fatigue^1^, mean (SD) | 85.2 (6.5)* | 85.4 (6.5)* | 85.3 (6.1)* | 85.8 (6.2)* | 85.4 (6.3)* |
| Cognitive functioning^1^, mean (SD) | 24.8 (11.9)* | 24.2 (11.4)* | 24.7 (11.4)* | 24.2 (11.4)* | 24.5 (11.5)* |
| Physical health (SF-36)^2^, mean (SD) | 90.8 (9.0)* | 90.9 (9.1)* | 91.4 (7.7)* | 90.7 (9.3)* | 91.0 (8.7)* |
| Mental health (SF-36)^2^, mean (SD) | 81.7 (12.1)* | 82.0 (12.1)* | 81.6 (12.7)* | 82.2 (11.6)* | 81.9 (12.2)* |
| Warm glow, N (%) | 478 (60.9)* | 552 (63.4)* | 609 (61.0)* | 449 (61.3)* | 2088 (61.7)* |
| Missing: * < 10%, ** 10-20%, *** > 20% | | | | | |
