## Supplemental table 3 for "Effectiveness of Ferritin-guided Donation Intervals in Blood Donors: Results of the Stepped-wedge Cluster Randomised FIND’EM Trial"

|  | | **Table S3.** Associations of implementation of ferritin-guided donation intervals with primary and secondary study outcomes. | | | | | | | | |
| --- | --- | --- | --- | --- | --- | --- | --- | --- | --- | --- |
|  |  | |  |  | Time since implementation | | | | | |
| Outcome | Group | |  | N | 6-11 months^1^ | 12-17 months | 18-23 months | 24-29 months | 30-35 months | 36+ months |
| Ferritin (ng/mL, log10) | PRE^2^ | | Coef (95% CI) | 9,683 | 0.01 (-0.02 – 0.04) | **0.06 (0.03 – 0.08)** | **0.07 (0.03 – 0.10)** | **0.09 (0.06 – 0.11)** | **0.10 (0.06 – 0.15)** | **0.14 (0.10 – 0.18)** |
|  | POST^3^ | |  | 6,534 | 0.04 (0.00 – 0.07) | **0.09 (0.07 – 0.12)** | **0.11 (0.08 – 0.15)** | **0.14 (0.11 – 0.17)** | **0.19 (0.15 – 0.24)** | **0.22 (0.18 – 0.25)** |
|  | Males | |  | 15,737 | **0.08 (0.05 – 0.10)** | **0.17 (0.15 – 0.19)** | **0.18 (0.16 – 0.21)** | **0.21 (0.19 – 0.24)** | **0.22 (0.19 – 0.26)** | **0.24 (0.22 – 0.27)** |
| Iron deficiency^4^ | PRE | | OR  (95% CI) | 9,683 | 1.08 (0.92 – 1.27) | **0.76 (0.67 – 0.86)** | **0.69 (0.57 – 0.82)** | **0.65 (0.56 – 0.77)** | **0.61 (0.47 – 0.79)** | **0.44 (0.34 – 0.55)** |
|  | POST | |  | 6,534 | 0.81 (0.64 – 1.01) | **0.59 (0.49 – 0.70)** | **0.45 (0.34 – 0.59)** | **0.41 (0.32 – 0.52)** | **0.24 (0.15 – 0.37)** | **0.19 (0.12 – 0.27)** |
|  | Males | |  | 15,737 | **0.55 (0.47 – 0.65)** | **0.29 (0.25 – 0.33)** | **0.23 (0.18 – 0.28)** | **0.16 (0.13 – 0.20)** | **0.15 (0.10 – 0.21)** | **0.19 (0.14 – 0.24)** |
| Hemoglobin (g/dL) | PRE | | Coef (95% CI) | 11,994 | 0.02 (-0.04 – 0.08) | **0.07 (0.03 – 0.12)** | **0.17 (0.11** **– 0.23)** | **0.16 (0.10 – 0.21)** | 0.03 (-0.05 – 0.10) | **0.17 (0.10 – 0.23)** |
|  | POST | |  | 7,683 | 0.09 (0.01 – 0.18) | 0.04 (-0.01 – 0.10) | **0.17 (0.09 – 0.25)** | **0.17 (0.10 – 0.24)** | 0.07 (-0.03 – 0.18) | **0.17 (0.08 – 0.25)** |
|  | Males | |  | 17,919 | **0.14 (0.08 – 0.20)** | **0.21 (0.17 – 0.25)** | **0.32 (0.26 – 0.37)** | **0.31 (0.26 – 0.37)** | **0.29 (0.21 – 0.36)** | **0.37 (0.31 – 0.43)** |
| Hemoglobin deferral^5^ | PRE | | OR  (95% CI) | 11,994 | 0.88 (0.67 – 1.15) | 0.81 (0.66 – 0.98) | **0.54 (0.39 – 0.74)** | **0.64 (0.49 – 0.83)** | **0.56 (0.37 – 0.81)** | 0.66 (0.48 – 0.90) |
|  | POST | |  | 7,683 | 0.89 (0.56 – 1.12) | 0.82 (0.59 – 1.12) | 0.64 (0.36 – 1.04) | **0.46 (0.27 – 0.73)** | 0.78 (0.41 – 1.37) | 0.49 (0.26 – 0.83) |
|  | Males | |  | 17,919 | 0.80 (0.55 – 1.12) | **0.50 (0.36 – 0.66)** | **0.35 (0.20 – 0.56)** | **0.44 (0.29 – 0.64)** | **0.23 (0.09 – 0.48)** | **0.23 (0.11 – 0.41)** |
| Donor return^6^ | PRE | | OR  (95% CI) | 11,994 | 0.90 (0.77 – 1.04) | **0.86 (0.78 – 0.96)** | 0.91 (0.78 – 1.05) | **0.76 (0.67 – 0.88)** | **0.77 (0.65 – 0.92)** | 0.88 (0.76 – 1.03) |
|  | POST | |  | 7,683 | 1.07 (0.88 – 1.31) | **0.81 (0.71 – 0.93)** | **0.73 (0.61 – 0.89)** | **0.72 (0.61– 0.85)** | 0.74 (0.59 – 0.94) | 0.77 (0.64 – 0.94) |
|  | Males | |  | 17,919 | 0.85 (0.73 – 0.98) | **0.86 (0.77 – 0.96)** | **0.74 (0.64 – 0.86)** | **0.67 (0.59 – 0.77)** | **0.64 (0.54 – 0.77)** | **0.56 (0.49 – 0.65)** |
| Restless legs syndrome | PRE | | OR (95% CI) | 2,140 | 1.58 (0.99 – 2.49) | 0.98 (0.57 – 1.65) | **2.23 (1.34 – 3.63)** | **2.27 (1.46 – 3.50)** |  |  |
|  | POST | |  | 1,303 | **2.20 (1.42 – 3.41)** | 0.54 (0.28 – 0.97) | 1.14 (0.53 – 2.23) | **2.27 (1.37 – 3.71)** |  |  |
|  | Males | |  | 3,316 | 1.09 (0.70 – 1.64) | 0.73 (0.45 – 1.14) | **1.80 (1.12 – 2.80)** | 1.49 (1.01 – 2.16) |  |  |
| Pica | PRE | | OR (95% CI) | 2,345 | 1.06 (0.29 – 2.18) | 0.75 (0.29 – 1.67) | 1.28 (0.50 – 2.88) | 0.80 (0.29 – 1.86) |  |  |
|  | POST | |  | 1,465 | 0.53 (0.08 – 2.05) | 1.66 (0.60 – 4.38) | 1.62 (0.25 – 6.31) | 1.93 (0.52 – 5.87) |  |  |
|  | Males | |  | 3,545 | 0.68 (0.32 – 1.31) | 1.00 (0.55 – 1.72) | 1.11 (0.48 – 2.25) | 0.91 (0.47 – 1.65) |  |  |
| Fatigue^7^ | PRE | | Coef (95% CI) | 2,360 | -0.33 (-1.05 – 0.39) | 0.71 (-0.03 – 1.44) | -0.86 (-1.75 – 0.04) | -0.38 (-1.17 – 0.42) |  |  |
|  | POST | |  | 1,471 | -0.29 (-1.09 – 0.51) | 0.32 (-0.45 – 1.09) | -0.16 (-1.31 – 1.00) | -0.49 (-1.54 – 0.55) |  |  |
|  | Males | |  | 3,558 | -0.40 (-0.92 – 0.12) | -0.50 (-1.01 – 0.01) | -0.78 (-1.61 – 0.04) | -0.27 (-0.82 – 0.28) |  |  |
| Cognitive functioning^7^ | PRE | | Coef (95% CI) | 2,342 | 0.14 (-1.35 – 1.64) | -1.46 (-2.94 – 0.03) | -0.92 (-2.62 – 0.77) | 0.87 (-0.82 – 2.56) |  |  |
|  | POST | |  | 1,464 | 0.87 (-0.77 – 2.51) | 1.08 (-0.50 – 2.67) | 2.59 (-0.06 – 5.23) | 0.86 (-1.12 – 2.84) |  |  |
|  | Males | |  | 3,534 | 0.05 (-1.04 – 1.13) | 0.81 (-0.29 – 1.90) | -0.59 (-2.14 – 0.96) | -0.24 (-1.38 – 0.90) |  |  |
| Physical health (SF-36)^8^ | PRE | | Coef (95% CI) | 2,398 | -0.04 (-0.76 – 0.68) | 0.32 (-0.39 – 1.04) | 0.11 (-0.79 – 1.00) | -0.64 (-1.47 – 0.19) |  |  |
|  | POST | |  | 1,468 | -0.33 (-1.32 – 0.66) | -1.12 (-2.14 – -0.09) | -1.01 (-2.57 – 0.54) | 0.78 (-0.43 – 1.98) |  |  |
|  | Males | |  | 3,582 | -0.43 (-0.98 – 0.12) | -0.47 (-1.00 – 0.07) | -0.35 (-1.07 – 0.37) | -0.12 (-0.64 – 0.40) |  |  |
| Mental health (SF-36)^8^ | PRE | | Coef (95% CI) | 2,404 | -1.25 (-2.25 – -0.24) | -0.04 (-1.01 – 0.92) | -1.29 (-2.90 – 0.32) | -0.98 (-2.14 – 0.18) |  |  |
|  | POST | |  | 1,476 | -0.25 (-1.20 – 0.79) | 0.30 (-0.75 – 1.34) | -1.37 (-3.16 – 0.42) | 0.06 (-1.13 – 1.24) |  |  |
|  | Males | |  | 3,595 | 0.10 (-.56 – 0.75) | -0.46 (-1.13 – 0.22) | -0.16 (-1.10 – 0.79) | 0.925 (-1.02 – 0.34) |  |  |
| Warm glow | PRE | | OR (95% CI) | 2,167 | 0.99 (0.77 – 1.26) | 1.41 (1.11 – 1.80) | 1.03 (0.75 – 1.40) | 1.06 (0.81 – 1.39) |  |  |
|  | POST | |  | 1,403 | 0.86 (0.63 – 1.18) | 0.80 (0.60 – 1.07) | 0.43 (0.27 – 0.68) | 0.90 (0.62 – 1.34) |  |  |
|  | Males | |  | 3,386 | 0.83 (0.67 – 1.02) | 0.97 (0.79 – 1.19) | 0.72 (0.55 – 0.95) | 0.81 (0.65 – 1.00) |  |  |
| Bold text indicates significance at p < 0.008 (Bonferroni correction: 0.05/6) for primary outcomes and p<0.01 (0.05/4) for secondary outcomes;  All analyses are scaled and adjusted for age, height, and weight ^1^ Reference group: no implementation and 0-5 months since implementation ^2^ PRE: Premenopausal females (age ≤ 48) ^3^ POST: Postmenopausal females (age > 48) ^4^ Ferritin < 15 ng/mL ^5^ For males ≥ 13.5 g/dL, for females ≥ 12.5 g/dL  ^6^ Within six months of the first allowed date for the next donation, additionally adjusted for whether return should have been during COVID pandemic ^7^ Higher score indicates worse result ^8^ Higher score indicates better result | | | | | | | | | | |
