## Supplemental table 4 for "Effectiveness of Ferritin-guided Donation Intervals in Blood Donors: Results of the Stepped-wedge Cluster Randomised FIND’EM Trial"

|  | | **Table S4.** Odds ratios of donor return within six months of the first allowed date for the next donation | | | | | | | |
| --- | --- | --- | --- | --- | --- | --- | --- | --- | --- |
|  |  | |  | Time since implementation | | | | | |
| Group |  | | N | 6-11 months^1^ | 12-17 months | 18-23 months | 24-29 months | 30-35 months | 36+ months |
| PRE^2^ | Ferritin > 30 | | 4,253 | 1.13 (0.88 – 1.45) | 1.02 (0.85 – 1.23) | 1.11 (0.84 – 1.45) | 1.01 (0.79 – 1.29) | 0.83 (0.59 – 1.17) | 0.97 (0.73 – 1.30) |
|  | Ferritin ≤ 30 | | 5,430 | **0.79 (0.65 – 0.97)** | 0.82 (0.70 – 0.96) | 0.91 (0.71 – 1.17) | **0.70 (0.55 – 0.87)** | 1.07 (0.77 – 1.50) | 1.00 (0.74 – 1.34) |
| POST^3^ | Ferritin > 30 | | 3,436 | 1.03 (0.77 – 1.39) | 1.02 (0.82 – 1.27) | 0.80 (0.59 – 1.11) | 0.82 (0.62 – 1.09) | 0.87 (0.61 – 1.24) | 0.92 (0.68 – 1.25) |
|  | Ferritin ≤ 30 | | 3,098 | 1.13 (0.84 – 1.53) | 0.76 (0.60 – 0.95) | 0.89 (0.63 – 1.27) | **0.59 (0.43 – 0.82)** | 0.99 (0.60 – 1.65) | 1.29 (0.81 – 2.08) |
| Males | Ferritin > 30 | | 9,166 | 0.99 (0.81 – 1.22) | 1.11 (0.96 – 1.29) | 0.87 (0.71 – 1.09) | 0.92 (0.76 – 1.11) | 0.81 (0.63 – 1.05) | 0.76 (0.61 – 0.93) |
|  | Ferritin ≤ 30 | | 6,571 | **0.59 (0.46 – 0.78)** | **0.70 (0.56 – 0.88)** | **0.53 (0.38– 0.73)** | **0.37 (0.27 – 0.50)** | 0.69 (0.43 – 1.14) | **0.35 (0.25 – 0.51)** |
| Bold text indicates significance at p < 0.008 (Bonferroni correction applied: 0.05/6); all analyses are adjusted for age, height, and weight ^1^ Reference group: no implementation or 0-5 months since implementation ^2^ PRE: Premenopausal females (age ≤ 48) ^3^ POST: Postmenopausal females (age > 48) | | | | | | | | | |
