## Supplemental table 5 for "Effectiveness of Ferritin-guided Donation Intervals in Blood Donors: Results of the Stepped-wedge Cluster Randomised FIND’EM Trial"

|  | | **Table S5.** Odds ratios of donor return within six months of the first allowed date for the next donation | | | | | | | |
| --- | --- | --- | --- | --- | --- | --- | --- | --- | --- |
|  |  | |  | Time since implementation | | | | | |
| Group |  | | N | 6-11 months^1^ | 12-17 months | 18-23 months | 24-29 months | 30-35 months | 36+ months |
| PRE^2^ | Hb deferred | | 812 | 0.76 (0.44 – 1.31) | 0.83 (0.56 – 1.26) | 0.70 (0.34 – 1.42) | 0.44 (0.23 – 0.83) | 1.12 (0.47 – 2.76) | 0.96 (0.47 – 1.97) |
|  | Extended interval | | 5,430 | 0.79 (0.65 – 0.97) | 0.82 (0.70 – 0.96) | 0.91 (0.71 – 1.17) | 0.70 (0.55 – 0.87) | 1.07 (0.77 – 1.50) | 0.77 (0.74 – 1.34) |
| POST^3^ | Hb deferred | | 299 | 0.34 (0.13 – 0.88) | **0.37 (0.18 – 0.77)** | 0.33 (0.09 – 1.15) | 0.67 (0.20 – 2.38) | 0.94 (0.22 – 4.93) | 0.62 (0.16 – 2.75) |
|  | Extended interval | | 3,098 | 1.13 (0.84 – 1.53) | 0.76 (0.60 – 0.95) | 0.89 (0.63 – 1.27) | **0.59 (0.43 – 0.82)** | 0.99 (0.60 – 1.65) | 1.29 (0.81 – 2.08) |
| Males | Hb deferred | | 451 | 0.78 (0.33 – 2.08) | 0.62 (0.30 – 1.35) | 0.55 (0.16 – 2.27) | **0.22 (0.08 – 0.60)** | 0.15 (0.02 – 0.93) | 0.74 (0.16 – 5.39) |
|  | Extended interval | | 6,571 | **0.59 (0.46 – 0.78)** | **0.70 (0.56 – 0.88)** | **0.53 (0.38 – 0.73)** | **0.37 (0.27 – 0.50)** | 0.69 (0.43 – 1.14) | **0.35 (0.25 – 0.51)** |
| Bold text indicates significance at p < 0.008 (Bonferroni correction applied: 0.05/6); all analyses are adjusted for age, height, and weight ^1^ Reference group: no implementation or 0-5 months since implementation ^2^ PRE: Premenopausal females (age ≤ 48) ^3^ POST: Postmenopausal females (age > 48) | | | | | | | | | |
