## Supplemental table 6 for "Effectiveness of Ferritin-guided Donation Intervals in Blood Donors: Results of the Stepped-wedge Cluster Randomised FIND’EM Trial"

|  | | **Table S6.** Odds ratios of donor return within six months of the first allowed date for the next donation by donation center type | | | | | | | |
| --- | --- | --- | --- | --- | --- | --- | --- | --- | --- |
|  |  | |  | Time since implementation | | | | | |
| Group |  | | N | 6-11 months^1^ | 12-17 months | 18-23 months | 24-29 months | 30-35 months | 36+ months |
| PRE^2^ | Fixed | | 11,027 | 0.83 (0.71 – 0.97) | 0.92 (0.82 – 1.04) | 0.99 (0.83 – 1.17) | 0.86 (0.73 – 1.01) | 0.96 (0.78 – 1.17) | 1.09 (0.91 – 1.30) |
|  | Mobile | | 967 | 1.67 (0.92 – 3.05) | 1.28 (0.90 – 1.82) | 0.80 (0.39 – 1.59) | 0.37 (0.20 – 0.68) | 1.00 (0.48 – 2.06) | 1.34 (0.71 – 2.53) |
| POST^3^ | Fixed | | 6617 | 0.95 (0.76 – 1.18) | 0.86 (0.72 – 1.02) | 0.80 (0.63 – 1.03) | 0.72 (0.58 – 0.90) | 0.83 (0.62 – 1.11) | 0.93 (0.73 – 1.18) |
|  | Mobile | | 1,066 | 1.25 (0.75 – 2.07) | 1.12 (0.80 – 1.57) | 0.75 (0.41 – 1.34) | 0.68 (0.40 – 1.17) | 2.24 (1.13 – 4.53) | 1.31 (0.70 – 2.44) |
| Males | Fixed | | 16223 | 0.87 (0.74 – 1.03) | 0.86 (0.76 – 0.98) | **0.72 (0.61 – 0.87)** | **0.67 (0.57 – 0.79)** | **0.66 (0.54 – 0.82)** | **0.59 (0.50 – 0.70)** |
|  | Mobile | | 1,696 | 0.66 (0.44 – 0.98) | 1.22 (0.91 – 1.64) | 1.02 (0.91 – 1.64) | 0.78 (0.51 – 1.20) | 1.19 (0.62 – 2.32) | 1.06 (0.65 – 1.76) |
| Bold text indicates significance at p < 0.008 (Bonferroni correction applied: 0.05/6); all analyses are adjusted for age, height, and weight ^1^ Reference group: no implementation or 0-5 months since implementation ^2^ PRE: Premenopausal females (age ≤ 48) ^3^ POST: Postmenopausal females (age > 48) | | | | | | | | | |
